# The use of computerised testing to assess cognitive performance in people with HIV in South Africa

**DOI:** 10.64898/2026.08.27.26361083

**Authors:** Evan C. Edmond, Anna J. Dreyer, Alan Winston, Saye H. Khoo, John Joska, Sam Nightingale

## Abstract

**Background:** Computerised cognitive testing may address the global challenge in identifying cognitive changes in people living with HIV scalably and affordably. We assessed a computerised battery (CB) of cognitive tests, in a prospective cohort (CONNECT) of people with HIV in a low-income peri-urban area of Cape Town, South Africa during a national programmatic switch from efavirenz-to dolutegravir-based antiretroviral therapy (ART).

**Methods:** We recruited 170 people with HIV and 91 people without HIV (controls) (140[82%] and 41[45%] followed up). The CB and gold-standard pen&paper cognitive testing (P&P) were performed at both timepoints. Technology familiarity/use questionnaire data were also collected. We compared performance in detecting lower group-level cognitive performance associated with efavirenz treatment. Furthermore, the CB was compared to P&P in classifying individuals with low cognitive performance, correlation of global test scores and domain-level scores between batteries, and practice effects between timepoints. Exploratory principal component analysis was also performed.

**Results:** People with HIV on efavirenz at baseline had lower performance on the computerised battery than controls, ΔT=2.6, p=0.0047. This difference was lost after switching to dolutegravir-based ART at follow-up. CB and P&P global T were moderately correlated (R^2^=0.203, p<0.001), and the CB performed moderately in classification of low cognitive performance against the gold standard (AUC 0.70, sensitivity 0.52, specificity 0.76, PPV 0.40, and NPV 0.84). Selecting the first three principal components improved both classification of low cognitive performance (AUC 0.77) and correlation strength with P&P global T (R^2^=0.3, p<0.001). The CB did not show practice effects. Most participants owned a mobile phone (95%, 85.9% of these smartphones). Performance was better in smartphone owners (ΔT=1.8) and computer owners (23%, ΔT=1.8).

**Conclusions:** Delivering computerised cognitive testing was feasible in this low-income southern African setting. The CB showed reasonable construct validity (detecting known lower cognitive performance associated with efavirenz-ART) and may detect broad cognitive characteristics such as processing speed and accuracy. However, correlation of CB results with gold standard P&P testing was low-moderate and may limit its applicability as a diagnostic tool. This might be improved by including a wider range of cognitive domains tested in the CB, or data driven analysis. Brief CBs may fulfil an initial screening role, followed by more detailed clinical assessment.

## Introduction

Cognitive testing is a key part of the clinical assessment to identify cognitive disorders. The traditional approach of neuropsychological pen & paper testing is richly informative but time consuming and dependent on availability, training, and funding of skilled testers. Expert availability per capita to diagnose and manage disorders of cognition is ∼100x greater in high income countries (2.2/100k) than in low-middle income countries (0.02/100k)^1^. This gap is not feasibly addressable by training new neuropsychology clinicians at current rates. The 2023 WHO dementia research blueprint^2^ advocates for novel clinical diagnostics that are applicable to diverse settings and the entire disease spectrum including prodromal disease.

Computerised cognitive testing could address this need, with various tools applied to screen for low cognitive performance in people with HIV in sub-Saharan Africa^3–5^. Validation concerns remain with regards to case/cohort characteristics, lack of control groups, demographic data, and most importantly – construct validity (“does the measure behave as though it is measuring the (indirectly measured) property”)^6,7^.

This study investigated one such tool – the Cogstate brief computerised battery (CB) – in the CONNECT study, a prospective cohort study of people with HIV in a low-income peri-urban area of Cape Town, South Africa, undertaken in the context of a national programmatic switch from efavirenz-to dolutegravir-based antiretroviral therapy (ART). Computerised cognitive testing has been used extensively in people with HIV, with most published work originating from North America^8,9^, Europe^10,11^, and Australia^12^, with one study in a low-income setting in Uganda^13^. Computerised cognitive testing may reduce the need for trained neuropsychometricians to administer and may yield significant process improvements when delivered in self-contained software packages that deliver stimuli, record responses, produce report data in a time-efficient, reliable, and reproducible manner.

We investigated computerised cognitive testing in a low-income cohort of South African people with and without HIV, by comparing performance on a brief CB with gold-standard neuropsychological pen & paper testing (P&P). First, we assessed whether the CB could detect a previously demonstrated effect (lower performance with efavirenz-based anti-retroviral treatment).

We compared CB vs. P&P performance head-to-head in classification of individuals with low cognitive performance, as well as correlating individuals’ scores between the two batteries. We further explored the raw CB data with principal component analysis aiming to infer what underlying cognitive features it might be measuring. We also assessed the appropriateness of a computerised interface in this population by collecting data on familiarity with technology and assessed the practice effects with <u>the CB</u> compared to P&P, i.e. changes in cognitive performance over time due to greater familiarity with the tests. Overall, this study assessed the feasibility of delivering a CB in this setting, as well as its construct validity.

## Methods

### Study design and participants

We recruited a prospective cohort of adults with and without HIV as part of the parent study: Cognition, Neuropsychiatric Symptoms and Neuroinflammation Switching to Dolutegravir in Cape Town (CONNECT), based at the Gugulethu Community Health Centre in a low-income peri-urban area of Cape Town, South Africa^14^.

Eligible people living with HIV were virally supressed with HIV-RNA below 1000 copies/ml, had been receiving efavirenz-based ART for at least 1 year, and were identified as eligible for switch to dolutegravir-based ART as part of the national programme. Individuals with factors that could confound cognitive testing were excluded, including: current substance use, high-risk or harmful alcohol use, history of central nervous system infection or major head injury, uncontrolled neurological conditions such as seizure disorders or established cerebrovascular disease, history of learning difficulty or severe intellectual disability, fewer than 7 years total education, or history of severe mental health disorder (schizophrenia, psychosis, or bipolar disorder). We excluded those with vertical HIV acquisition, currently being investigated or treated for active intercurrent illness such as infection or carcinoma, currently receiving treatment for tuberculosis, known or suspected to be pregnant, not fluent in English or isiXhosa, or with a contraindication to lumbar puncture. Full inclusion and exclusion criteria are described in previous work^14,15^.

People without HIV were recruited from the friends, relatives, and associates of people attending the Gugulethu HIV clinic, aiming for a similar sociodemographic background. Negative HIV status was confirmed by rapid test. People without HIV had the same exclusion criteria as people with HIV and were matched with people with HIV by age band and self-identified gender.

The study was approved by the University of Cape Town Faculty of Health Sciences Human Research Ethics Committee (017/2019). Written informed consent was obtained in the language of participant preference (English or isiXhosa). The study data relevant to this analysis (CB, P&P, demographic and technology use questionnaire) were accessed for analysis on 1^st^ Oct 2023 for research purposes. This information did not allow identification of individual participants during or after data collection.

### Technology use questionnaire

To assess the feasibility of computerised testing in a low-middle income setting, data were collected on participants’ computer and mobile phone ownership and usage, as well as overall familiarity with technology. Additional questions assessed difficulty with using the test laptop, as well as difficulty with each task. The full questionnaire is included in S1 Fig.

### Computerised cognitive test battery

Six tasks were selected from the Cogstate battery (Detection, Identification, One-back, Two-back, One card learning, and Set shifting). The test battery was presented on a laptop computer. The participant was shown how to use specified keys to respond “Yes” or “No” for each task. Full details of each task are given in Supplementary Materials. Participants underwent the full battery as a practice before the recorded test at baseline. No practice test was performed at follow up. Each task reports the following results: speed (mean reaction time for correct responses in milliseconds, log10 transformed), accuracy (arcsine transformation of the square root of the proportion of correct responses), and variability (log10 transformed standard deviation of response times). The principal outcome measures advised in the Cogstate literature are summarised in Table 1.

**Table 1.** Computerised battery task names, with corresponding principal outcome measures (speed – [mean reaction time for correct responses in milliseconds, log10 transformed], accuracy [arcsine transformation of the square root of the proportion of correct responses]) and corresponding cognitive domains.

| Task name | Principal outcome measure | Cognitive domain assessed |
| --- | --- | --- |
| Detection | speed | Processing speed |
| Identification | speed | Attention |
| One card learning | speed, accuracy | Visual learning |
| One-back, Two-back | accuracy | Working memory |
| Set shifting | accuracy | Executive function |

### Pen & Paper cognitive testing

Comprehensive cognitive testing was taken as our gold standard. This involved a standard battery of validated tests that assess seven cognitive domains, taking around 2 hours to administer. Tests were administered in either English or isiXhosa by a bilingual neuropsychology technician. A registered clinical neuropsychologist (AJD) supervised test administration and scoring protocols.

The cognitive domains, tests, and outcome variables were: (1) executive functioning, Color Trails Test 2 (CTT2) – completion time (in seconds); Wisconsin Card Sorting Test (WCST) – total score; (2) verbal learning and memory, Hopkins Verbal Learning Test-Revised (HVLT-R) – total across the three immediate recall trials, total on the delayed recall trial; (3) visuospatial learning and memory, Brief Visuospatial Memory Test-Revised (BVMT-R) – total across the immediate recall trials, total on the delayed recall trial; (4) verbal fluency, category fluency test – total number of animals / total number of fruits and vegetables named in 1 minute; (5) attention/working memory, Wechsler Adult Intelligence Scale-Third Edition (WAIS-III) Digit Span subtest – total raw score; (6) processing speed, CTT1 – completion time (in seconds); WAIS-III Digit Symbol Coding subtest – total raw score; WAIS-III Symbol Search – total raw score; (7) motor skills, Grooved Pegboard Test (GPT) non-dominant hand (NDH) – completion time (in seconds); Finger Tapping Test NDH – completion time (in seconds).

### Analysis

#### Global deficit scores

Raw scores for each task (both CB and P&P) were T-transformed (mean = 50, standard deviation = 10). As sex, years of education, and age were significantly associated with cognitive test performance in people without HIV – demographically corrected scores were calculated using standard regression-based norming processes^16,17^. Details of these methods have been described previously^18^. Domain T-scores for P&P testing were calculated by taking the average of T-scores of the tasks associated with each domain. Global T-scores were calculated by taking the average of the domain T-scores for P&P, and the principal outcome measure (Table 1) T-scores for the CB.

Global deficit scores were calculated based on transforming T scores (task T scores for CB, domain T scores for P&P) into deficit scores based on previously published thresholds^19^. Low cognitive performance on the CB was defined has having a mean deficit score greater than or equal to 0.5.

#### Group level comparison in relation to efavirenz-based ART switch

Independent samples t-tests were done to compare CB global T scores for people with HIV and without HIV (controls) at both baseline and follow-up after treatment switch. Mann-Whitney U tests were done at each timepoint to compare GDS classification of cognitive performance (low vs. normal/high) between groups.

#### Correlating CB and P&P results

The global T score derived from CB and P&P for each individual were plotted as a scatterplot, with least squares linear regression performed, and Pearson correlation coefficient calculated. Individual CB task scores were cross-correlated with individual P&P task scores by calculating Pearson correlation coefficients. Individual CB task scores were also cross-correlated with P&P testing derived cognitive domains by calculating Pearson correlation coefficients. The Pearson correlation coefficients were plotted in heatmaps/correlation matrices.

### Principal component analysis

To use the entire CB raw dataset, including speed, accuracy, and variability metrics from all CB tasks, we performed principal component analysis performed using the scikit-learn python toolkit (v1.4.2). Raw data for each input cognitive test was z-transformed (subtracting the mean and scaling to standard deviation) before PCA was performed using the sklearn.decomposition.PCA function. The cumulative explained variance was plotted in a screeplot. The same was repeated for P&P tasks to allow comparison of variance between the two batteries.

Similar proportions of explained variance were seen for both CB and P&P with 70% of variance explained by the first 6 components, so these were selected for plotting. The loadings (eigenvectors) of input features (individual tasks) onto the principal components were plotted as a heatmap.

We tested each principal component’s performance in classification of low/high cognitive performance against P&P by plotting ROC curves and summed those components that showed positive area under curve (AUC) [first three components] into a composite PCA derived metric of cognitive performance.

We also cross correlated the composite PCA derived metric with the P&P global T score in a scatterplot, performed least squares linear regression, and calculated the Pearson correlation coefficient.

### Technology use questionnaire

Independent samples t-tests were performed between the distributions of global CB T scores for questions where there were two responses (i.e. Yes/No), while for those questions with multiple responses, Kruskal-Wallis tests were performed.

### Practice effects

The difference in T score for each CB or P&P task for each individual between timepoints was calculated and averaged for each task. Paired t-test was performed to compare the distributions of T scores at each time point for each task.

## Results

One hundred and seventy people with HIV and 91 people without HIV were recruited between Aug 12, 2019, and Sept 16, 2022. 78.2% were female and mean age was 40.1 years.One hundred and forty people with HIV (82%) were followed up and 41 people without HIV (45%). The mean interval between baseline and follow-up was 342 days (SD 105) for people with HIV, and 365.3 days (SD 115.4) for people without HIV. Demographic data are described in Table 2.

**Table 2.** Baseline demographics for CONNECT cohort. Currency data collected in South African Rand is shown at a nominal conversion rate of R16.667 to 1 US dollar.

|  |  | Missing | Overall |
| --- | --- | --- | --- |
| n |  |  | 261 |
| Age at consent / years, mean (SD) |  |  | 40.1 (8.2) |
| Ethnicity, n (%) | Black South African | 1 | 260 (100.0) |
| Gender, n (%) | Female |  | 204 (78.2) |
|  | Male |  | 57 (21.8) |
| First language, n (%) | Afrikaans |  | 2 (0.8) |
|  | English |  | 7 (2.7) |
|  | Setswane |  | 1 (0.4) |
|  | Shona / isiShona |  | 5 (1.9) |
|  | Sotho |  | 4 (1.5) |
|  | Xhosa / isiXhosa |  | 241 (92.3) |
|  | Zulu / IsiZulu |  | 1 (0.4) |
| Testing language, n (%) | English |  | 28 (10.7) |
|  | Xhosa / isiXhosa |  | 233 (89.3) |
| Income level per month, n (%) | R0-R499 (\$0-\$29.99) | 10 | 40 (15.9) |
| | R500-R999 (\$30-\$59.99) | | 30 (12.0) |
| | R1000-R1999 (\$60-\$119.99) | | 59 (23.5) |
| | R2000-R2999 (\$120-\$179.99) | | 30 (12.0) |
| | R3000-R3999 (\$180-\$239.99) | | 30 (12.0) |
| | R4000-R4999 (\$240-\$299.99) | | 27 (10.8) |
| | R5000-R9999 (\$300-\$599.99) | | 35 (13.9) |
| HIV status, n (%) | Negative |  | 91 (34.9) |
|  | Positive |  | 170 (65.1) |

### Can computerised testing detect change associated with efavirenz to dolutegravir switch?

We found lower cognitive performance (CB global T score) in people with HIV at baseline compared to controls (Figs 1a and 1b). At follow-up, after the switch to dolutegravir-based ART, this difference was not seen. The overall pattern parallels previous work – people with HIV on efavirenz had lower performance than people without HIV at baseline, but both groups were similar at follow up after the treatment change^14^.

**Fig 1a.**
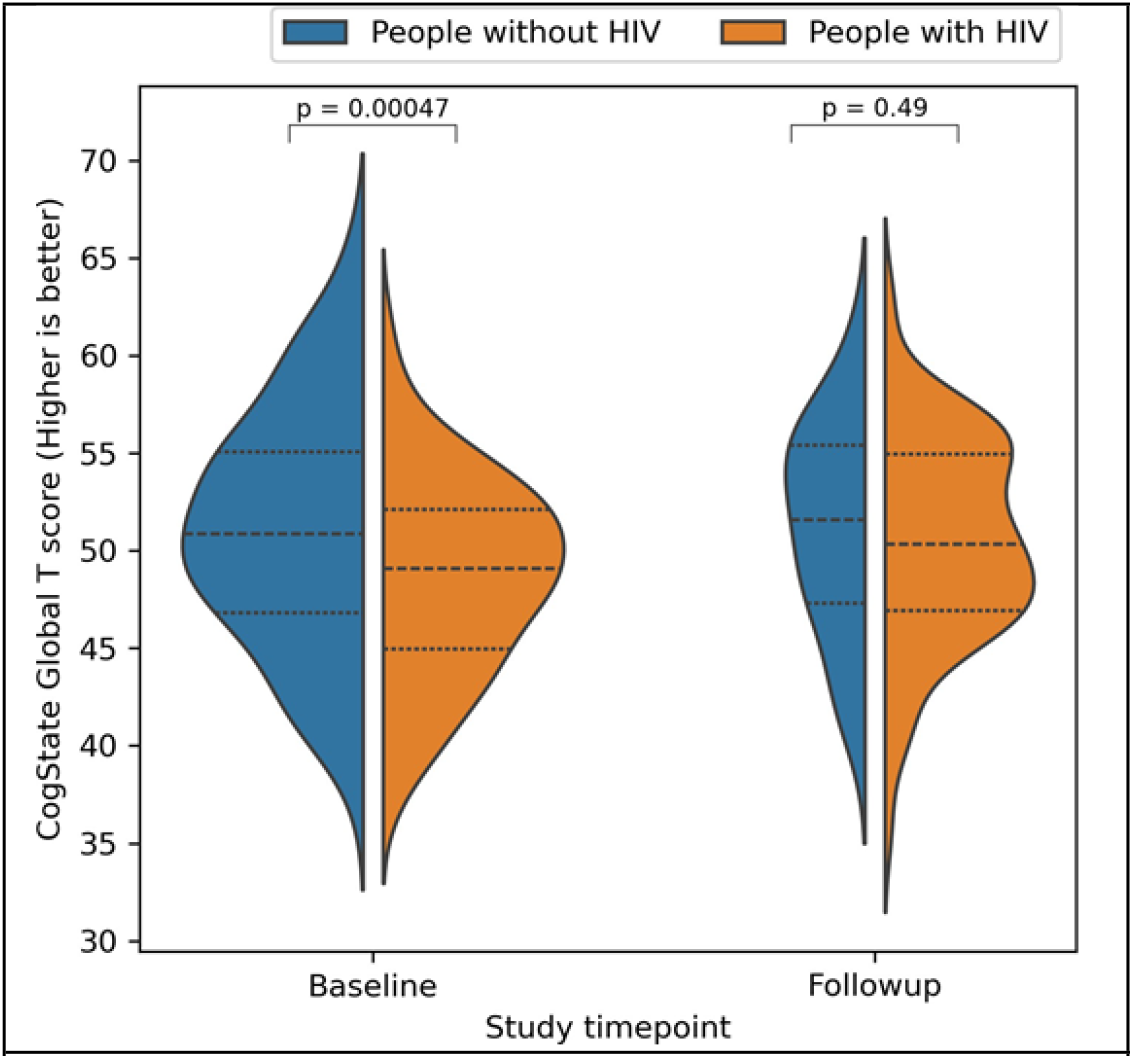
Violin plot of CB global T score split by HIV status and timepoint. The area of each violin is proportional to the number of participants. Dotted lines indicate quartiles.

**Fig 1b.**
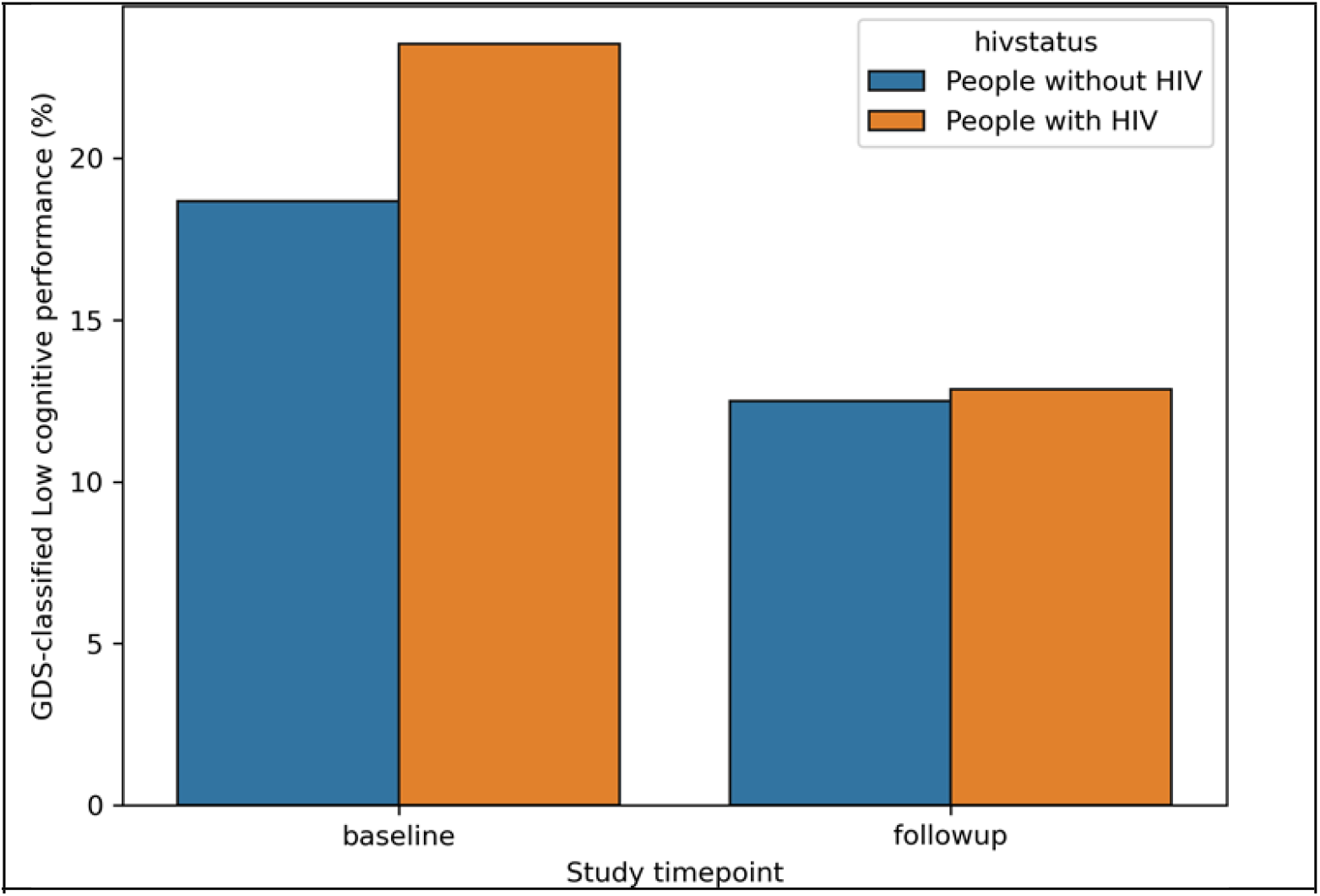
Percentage of individuals classified as “low cognitive performance” by GDS analysis of CB data.

### Global deficit scores

GDS generated using the computerised battery (CB) and P&P are cross tabulated in Table 3. This showed only moderate agreement between CB versus P&P testing. 30 (11.5%) participants were misclassified as cognitively impaired by CB, compared to the pen & paper testing gold standard. CB GDS classification had sensitivity 0.52, specificity 0.76, positive predictive value 0.40, and negative predictive value of 0.84 against the pen & paper testing gold standard. CB GDS performed best as a predictor of normal/high cognitive performance, with negative predictive value of 0.84.

**Table 3.** Classification of low cognitive performance (GDS) for computerised battery vs. Pen & Paper testing.

|  |  | Pen & Paper cognitive performance |  |
| --- | --- | --- | --- |
|  |  | Low | Normal/High |
| Computerised battery cognitive performance | Low | 32 | 48 |
|  | Normal/High | 30 | 152 |

The performance of the global CB T score against P&P in classification of cognitive performance is summarised in a receiver operating characteristic (ROC) curve in Fig 2a. Area-under-curve (AUC) of 0.70 suggests moderate performance of the global CB T score in identifying low cognitive performance.Plotting the CB global T score (primary outcome measures) against P&P global T score showed a moderate positive correlation at both baseline (Pearson R^2^ = 0.203, p < 0.001) and follow-up (Pearson R^2^ = 0.244, p < 0.001) timepoints (Figs 2b and 2c).

**Fig 2a.**
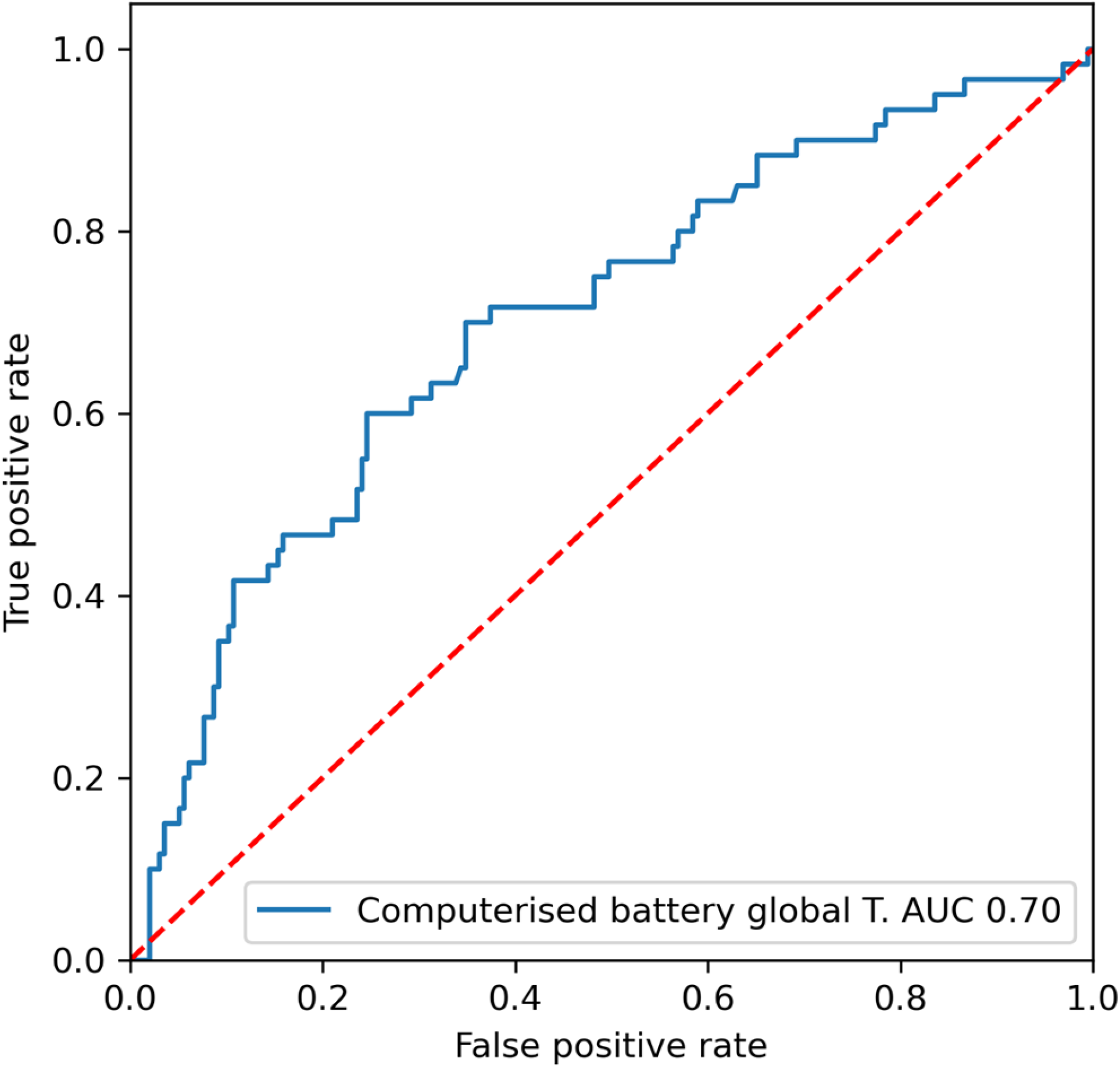
Receiver operating characteristic curve showing performance of computerised battery (CB) global T score against Pen & Paper based GDS classification of cognitive performance (at baseline).

**Fig 2b.**
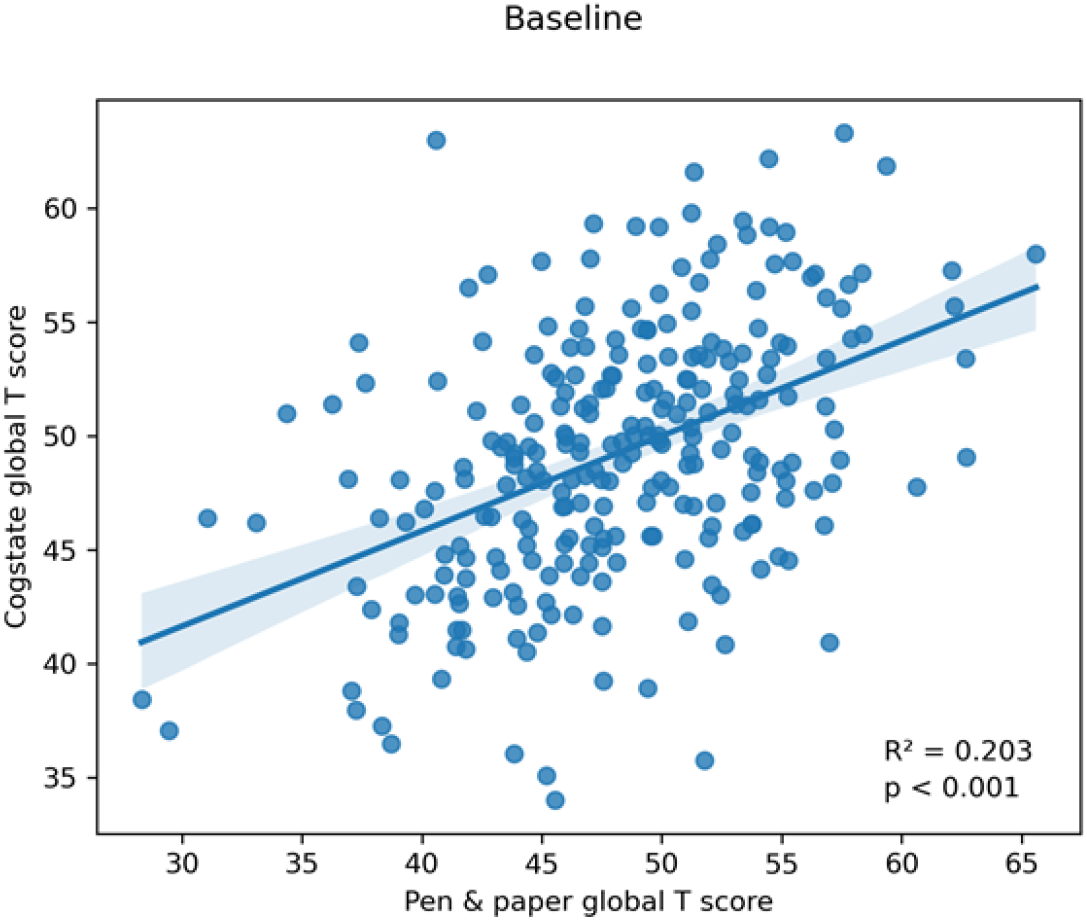
Linear regression of CB versus Pen & Paper global T scores at baseline.

**Fig 2c.**
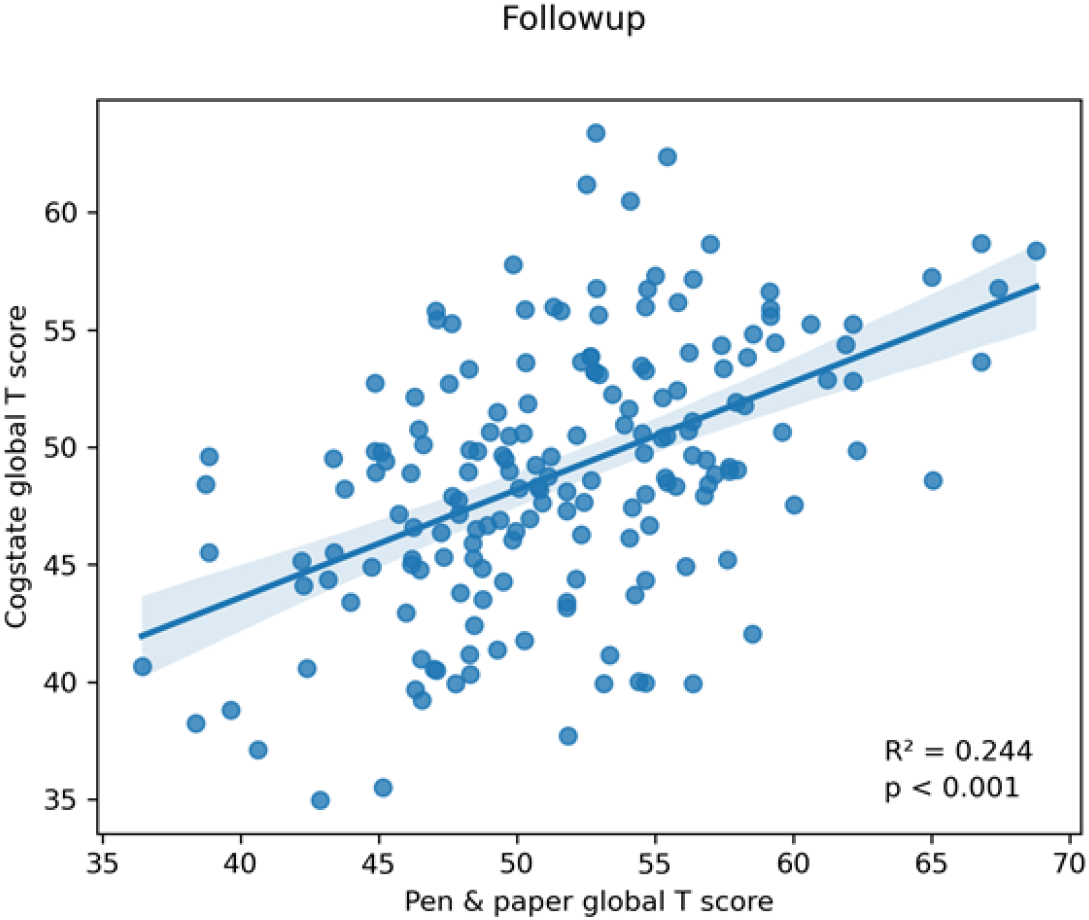
Linear regression of CB versus Pen & Paper global T scores at follow-up.

### Cross-correlation of CB principal outcomes with P&P domains

The principal CB outcome measures showed weak to moderate correlation with P&P domain scores (maximum Pearson R = 0.53 between CB One-back accuracy [Working Memory] and P&P Processing speed, Fig 3).Full cross-correlation matrices between raw CB scores (Speed, Variability, and Accuracy) vs. individual P&P tests and P&P derived cognitive domains are provided in S2 and S3 Figs.

**Fig 3:**
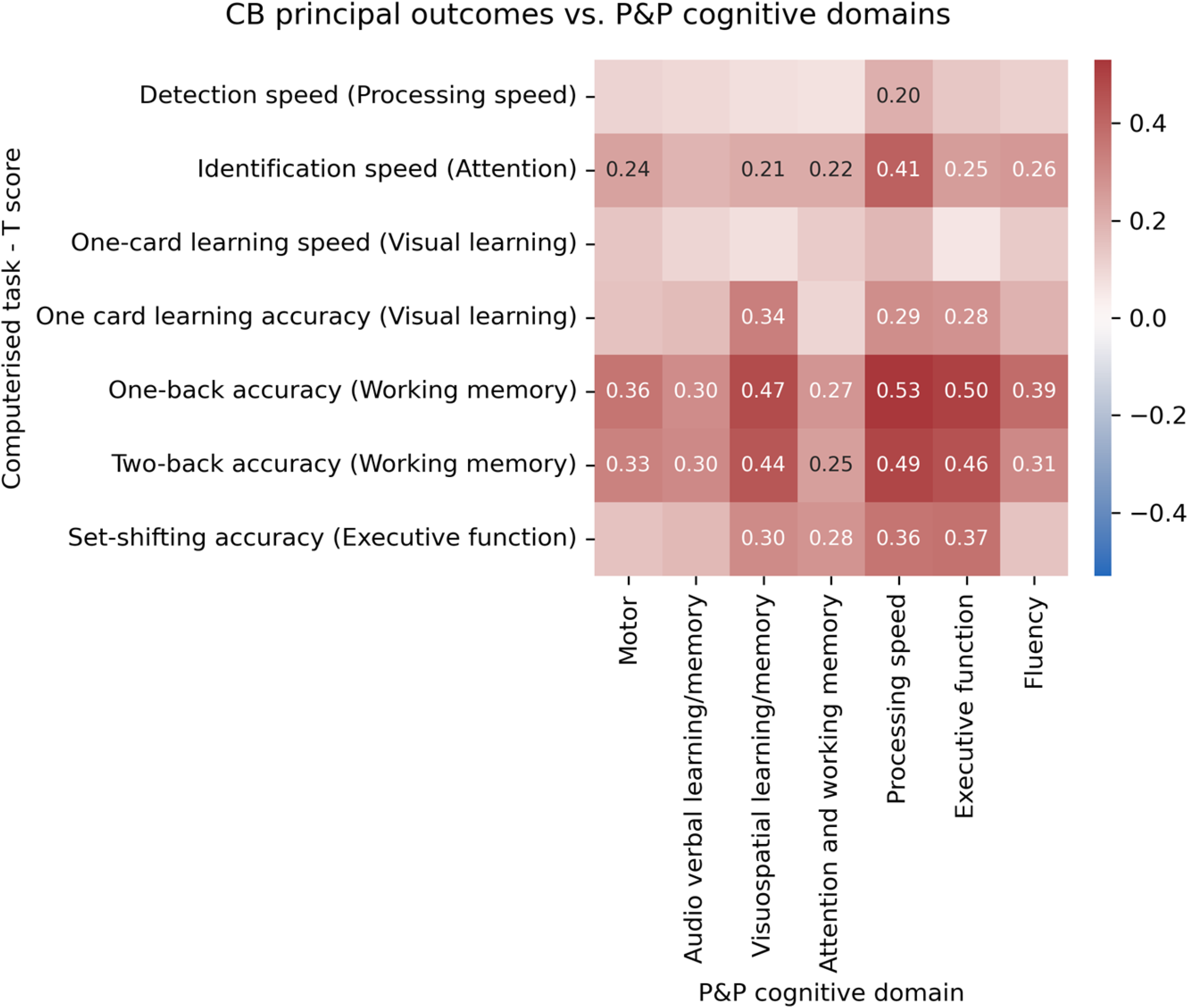
Heatmap of Pearson correlation coefficients between principal CB outcome measures vs. P&P cognitive domains.

### Data exploration with PCA

The loadings (eigenvectors) of the principal components derived from CB task scores are plotted in Fig 4, showing which input data features contributed the greatest variance for each component.

**Fig 4.**
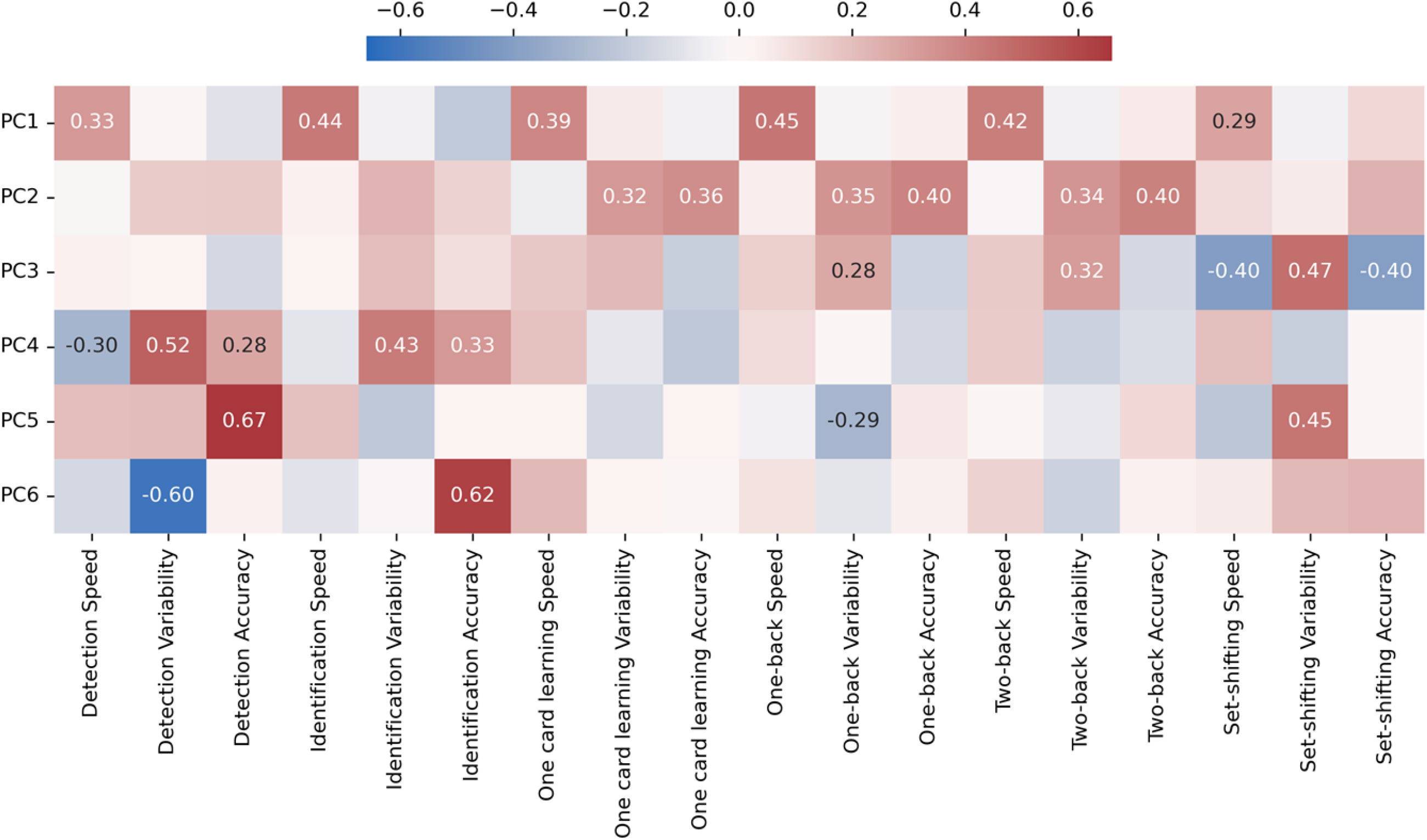
Heatmap of eigenvectors for each principal component.

#### PC1 – Speed

The first principal component is loaded onto the “Speed” metric (mean reaction time for correct responses in milliseconds, log10 transformed) from all CB tasks – this component likely captures a global “Processing speed”.

#### PC2 – Accuracy/Precision

The second component is loaded onto the “Variability” and “Accuracy” metrics from several tests (One-card learning, One-back and Two-back), with loading in the opposite direction onto the “Speed” metric. This likely captures a common “Precision” across multiple tasks.

#### PC3 – Executive function / Choice

The third component is most strongly loaded onto all three parameters from the Set shifting task, as well as “Variability” in the One-back and Two-back tasks. This likely represents a form of trade-off between speed/accuracy and variability in these tests and may reflect a common metric of “Executive function”.

#### PC4 – Precision-Speed trade-off

The fourth component shows a similar pattern as PC2 and is most strongly loaded in opposite directions onto Detection task “Speed” vs. Detection and Identification “Variability” and “Accuracy”. This may represent a trade-off between speed and precision with these two, predominantly processing speed based, tasks.

#### PC5/PC6 – Other variability/accuracy trade-offs? Noise?

PC5 and PC6 show strong loading onto a smaller number of metrics from disparate tasks, involving opposite-direction loadings onto “Variability” and “Accuracy” metrics. These do not suggest a clear or intuitive explanation and may reflect some other kind of trade-off between variability and accuracy in performing these tasks. They represent a small proportion of explained variability in these tasks and alternatively may reflect noise.

When using a combined metric derived from the first three principal components, the area under the ROC curve for classification of low cognitive performance improved to 0.77 (Fig 5a). and the strength of correlation with P&P global T score improved to Pearson R^2^ = 0.292 (Fig 5b).

**Fig 5a.**
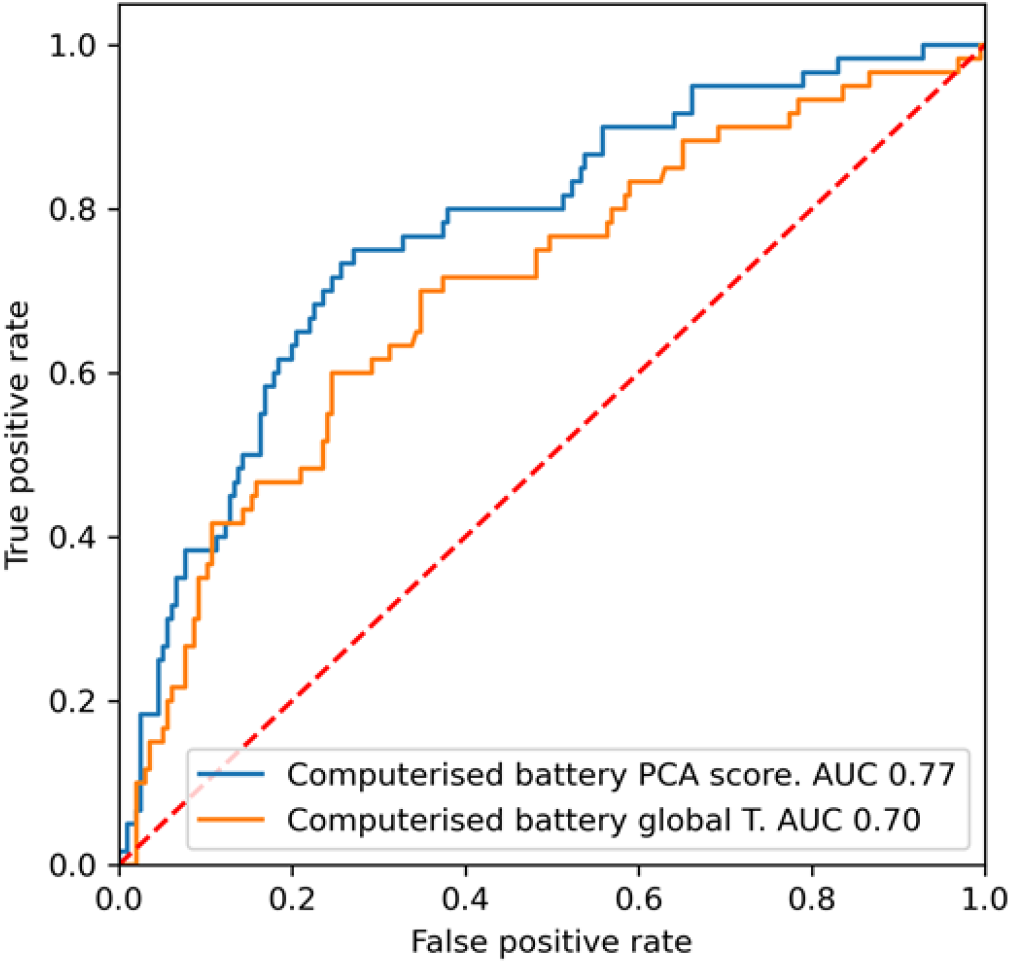
Receiver operating characteristic curve showing performance of first three CB principal components against P&P GDS classification of cognitive performance.

**Fig 5b.**
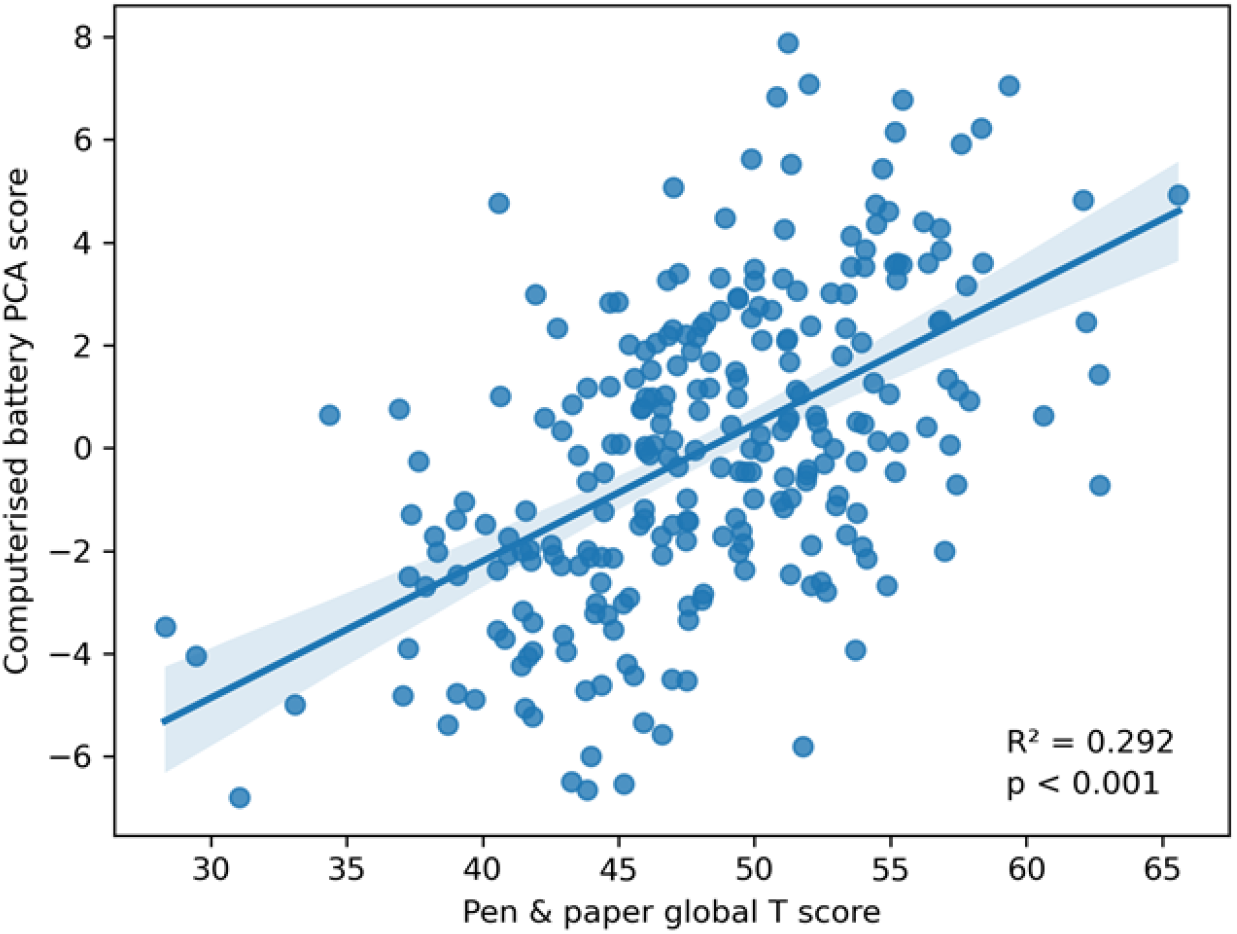
Scatter plot of first three CB principal components vs. P&P global T scores, with linear regression trendline, and 95% confidence interval of the regression estimate.

### Technology use questionnaire

The majority (77%) of participants did not own a computer, but mobile phone ownership was near universal (95%) – most of these (86%) with touchscreens. Most participants (79%) reported feeling somewhat comfortable, comfortable, or very comfortable using a computer. Most participants (85%) found it somewhat easy, easy, or very easy to use the computer during the testing session.

Participants who owned a computer had slightly better global CB performance (p = 0.014, effect size ΔT = 1.8). Those who owned a touchscreen mobile phone performed slightly better than those who owned a mobile phone without a touchscreen (p = 0.034, effect size ΔT = 1.8). Questionnaire data are presented in Table 4.

**Table 4.**
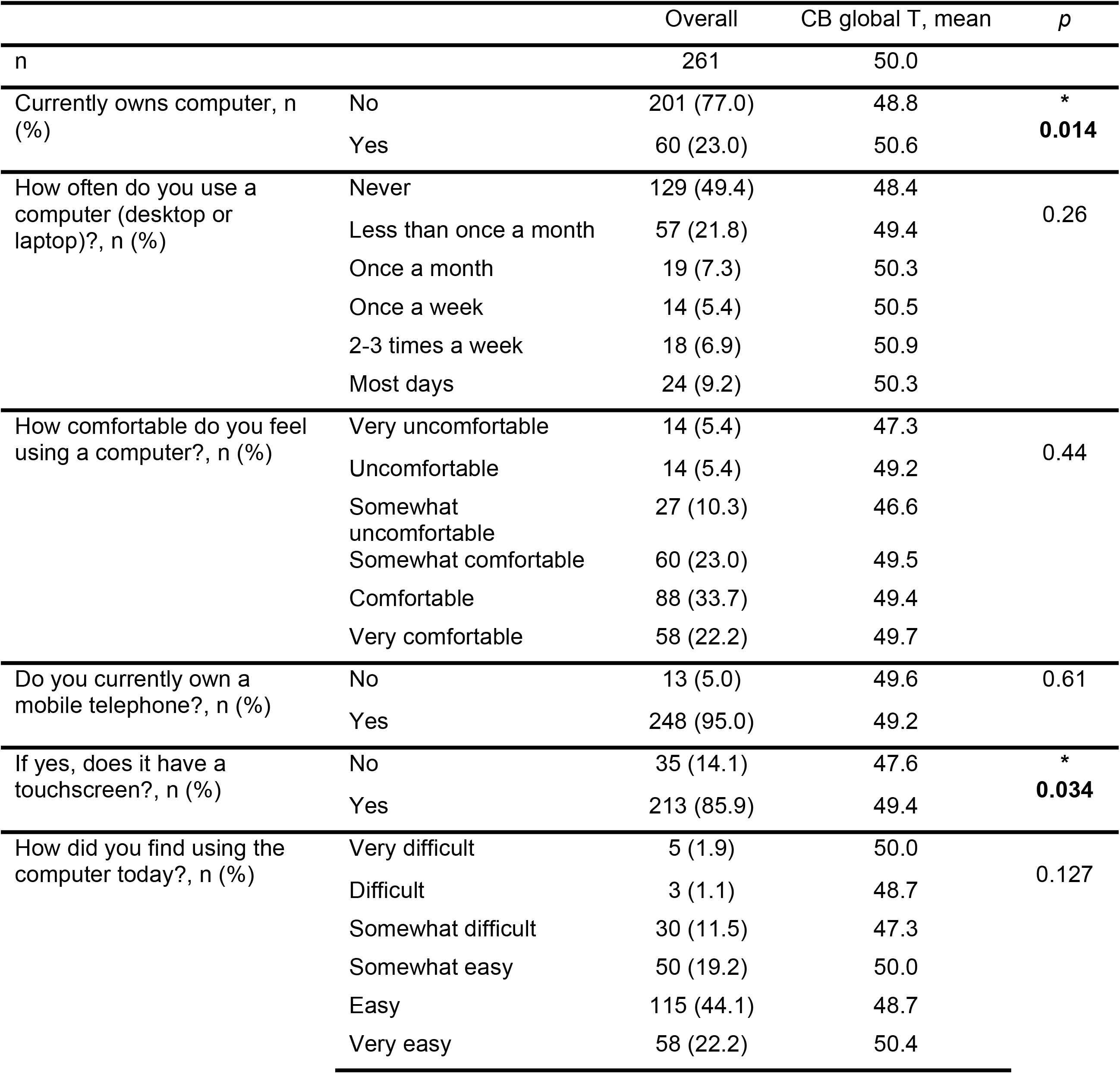
Results of technology use questionnaire.

### Practice effects – How does CB practice effect compare to P&P?

We found no significant between-session practice effects with CB tests (Table 5). Several pen & paper tests showed significant between-session improvement from baseline to follow-up, likely a practice effect (Table 6). There was a small global practice effect in the P&P global T score (ΔT = 2.57), but no significant effect was found with the CB global T score.

**Table 5.** Practice effects with computerised battery tasks. Positive delta (red highlight) means better performance at follow-up. Negative delta (blue highlight) means worse performance at follow-up.

| Task | Mean delta T<br>(T Visit 2 – T Visit 1) | p |
| --- | --- | --- |
| Detection speed (Processing speed) | -3.11 | 0.95 |
| Identification speed (Attention) | -0.14 | 0.54 |
| One-card learning speed (Visual learning) | -0.64 | 0.66 |
| One-card learning accuracy (Visual learning) | 2.59 | 0.07 |
| One-back accuracy (Working memory) | -0.98 | 0.76 |
| Two-back accuracy (Working memory) | 1.40 | 0.11 |
| Set-shifting accuracy (Executive function) | -1.29 | 0.75 |
| Computerised battery Global T | 0.14 | 0.43 |

**Table 6.** Practice effects with pen & paper tests. Positive delta (red highlight) means better performance at follow-up. Negative delta (blue highlight) means worse performance at follow-up.

| Task | Mean delta T<br>(T Visit 2 – T Visit 1) | p |
| --- | --- | --- |
| Grooved Pegboard Test | 4.33 | *0.01 |
| Finger Tapping test | -1.36 | 0.80 |
| HVLT-R - Total learning | 5.02 | *<0.01 |
| HVLT-R - Delayed recall | 2.76 | *0.05 |
| BVMT-R - Total learning | 4.16 | *<0.01 |
| BVMT-R - Delayed recall | 1.77 | 0.09 |
| Digit span | 3.78 | *0.01 |
| Digit symbol | 2.85 | *0.02 |
| Symbol search | 0.14 | 0.47 |
| CTT1 - Completion time | 2.39 | 0.09 |
| CTT2 - Completion time | 2.19 | 0.08 |
| WCST - total score | 2.77 | *0.05 |
| Fluency - animals | 1.58 | 0.20 |
| Fluency - fruit & veg | 2.30 | 0.07 |
| Global T | 2.57 | *<0.01 |

## Discussion

We found that delivering cognitive testing via the Cogstate computerised assessment was feasible in this cultural and sociodemographic context. The computerised battery (CB) was able to show the overall pattern of cognitive change, as demonstrated by gold standard testing published separately^14^, i.e. baseline differences between people with and without HIV that were not present following switch from efavirenz-to dolutegravir-based ART. The CB achieved this despite a shorter test duration without the requirement for neuropsychometric expertise, supporting that it could be a scalable approach to cognitive assessment in low-resource settings. We also showed that the CB had fewer practice effects than traditional cognitive testing (albeit with a full practice run before baseline testing) supporting its use for longitudinal assessment.

Most participants in this study had little access to computers, although the majority owned a mobile phone and most of these with touchscreens. In itself this is an important finding – touchscreen smartphones are globally ubiquitous even in low-income settings. Although we found slightly better global CB performance in participants who owned a computer or a smartphone with a touchscreen, this association was modest, and technology familiarity did not seem to be a major confound to the use of the CB in this setting. One explanation is that participants with greater familiarity with technology may have been more comfortable with the computerised interface of the CB and hence performed better. Another explanation is that technology ownership was a surrogate for higher socioeconomic status. Previous analysis of this cohort has demonstrated an association of cognitive performance with psychosocial confounds^15^.

However, the CB showed only modest associations with the results of pen & paper tests. If the CB had been used for classification in this study, 48 people would have been incorrectly labelled with low cognitive performance and 30 falsely reassured of normal cognitive performance. This represents 30% of the cohort being misclassified as compared with the gold standard. This may, in part, reflect the limitation of dichotomising a continuous variable in this way. There was a relatively small effect size found in lower cognitive performance in people living with HIV at baseline compared to the control group (T score difference of ∼3). This small difference may reflect the subtle neurotoxic effects of efavirenz, given that no group differences were evident at follow up^14^. In the modern era of effective ART, cognitive outcomes have improved and describing smaller cognitive differences by measuring the full spectrum of cognitive performance, rather than dichotomising to normal/abnormal, may be preferable, as proposed elsewhere^20^. However, when analysed this way, the correlation between the CB and P&P global T-score was moderate. The principal components derived from the CB dataset further show correlated variance along broad themes – *Speed*, *Accuracy*, and *Speed-Accuracy trade-offs* suggesting that coarse underlying cognitive features are being assessed.

There was a high false classification rate in our study – a proportion of our control group of people without HIV were classified as lower cognitive performance, both by the CB and by P&P. The group of people without HIV had all confounders excluded and hence were, by definition, cognitively unimpaired. A high false positive rate has been described in other neuro-HIV studies^21–25^. These findings support the arguments made in other papers^26^, including a recent international consensus statement^20^, that the results from testing of cognitive performance should not be interpreted in isolation to inform a classification of cognitive impairment, and require clinical context. To do so would risk misdiagnosis causing unnecessary anxiety and inappropriate investigation. An alternative use for the CB could be as screening tool, triggering more complete clinical assessment if positive. The moderate negative predictive value supports its use as a rule-out screen. However, low performance on CB testing should be interpreted very cautiously given the low sensitivity and specificity compared with gold standard^26^. There are now a wide range of computerised cognitive testing tools, including emerging artificial intelligence tools with a more naturalistic/conversational interface.

This study has several limitations. Most importantly, we used a relatively restricted set of Cogstate tasks in the brief battery. A more complete set of tasks involving more domains may have demonstrated a closer correlation with our gold standard. Our sample had well controlled HIV, and testing may have been limited by a low prevalence of cognitive impairment as described above. However, this cohort reflects the cognitive profile of people with HIV in our setting, where most are virally supressed on first line ART. The differential practice effects could be explained the by the inclusion of a full practice run for the CB, which was impractical for pen & paper testing. As no clinical assessments were performed on our cohort, a diagnosis of cognitive impairment cannot be made, and our gold standard reflects a surrogate outcome that may be inaccurate. This reflects a more general limitation of quantitative neuropsychometry, where the criterion validity of a given test battery in detecting the ground truth cognitive performance is limited. In future, integration of multimodal biomarkers from blood^27^, CSF^28^, and neurophysiology^29,30^ may facilitate early clinical diagnosis of neurodegenerative disorders in general and contextualise cognitive performance.

In summary, we found that computerised testing was feasible in this setting and was only minimally affected by familiarity with technology. A brief computerised battery has reasonable construct validity (consistent over time, detects known cognitive effects) and may be used to characterise broad cognitive characteristics. However, the weak correlation between CB and P&P results limits its use as a diagnostic tool and underlines the importance of a clinical assessment to categorise cognitive impairment.

## Data Availability

All data produced are available online on the Dryad data publishing platform doi: 10.5061/dryad.hx3ffbgvn

https://doi.org/10.5061/dryad.hx3ffbgvn

## Supplementary materials

### Methods

#### Computerised battery tasks

The **Detection** task aims to measure processing speed. The on-screen instructions read: “Has the card turned face up?”. A playing card is shown face down and flips to face up at a random interval. The participant is instructed to press “Yes” as soon as the card flips over, working as quickly and accurately as possible. Button presses before the card is flipped are marked as errors. The test continues until 35 correct responses are made, or 3 minutes are up.

The **Identification** task aims to measure attention using a choice reaction time task. The on-screen instructions read: “Is the card red?”. A playing card is shown face down and flips to face up. The participant is instructed to decide whether the card is red or not, and press “Yes” if it is red, and “No” if not, working as quickly and accurately as possible.

The **One-Back** task aims to measure working memory. The on-screen instructions read: “Is the previous card the same?”. A sequence of playing cards is shown, one at a time, face-up, in the centre of the screen. The participant is instructed to decide whether the card is the same as the previous card, pressing “Yes” if it is the same, and “No” if not, working as quickly and accurately as possible.

The **One Card Learning** task aims to measure visual learning. The on- screen instructions read: “Have you seen this card before in this test?”. A sequence of playing cards is shown, one at a time, face-up, in the centre of the screen. The participant is instructed to decide whether they have previously seen this card at any point in this task, pressing “Yes” if so, and “No” if not, working as quickly and accurately as possible. After the participant responds, the next card in the sequence is shown.

The **Two-Back** task aims to measure working memory. The on-screen instructions read: “Is the card the same as that shown two cards ago?”. A sequence of playing cards is shown, one at a time, face-up, in the centre of the screen. The participant is instructed to decide whether the card is the same as the card shown two cards previously, pressing “Yes” if it is the same, and “No” if not, working as quickly and accurately as possible. After the participant responds, the next card in the sequence is shown.

The **Set Shifting** task aims to measure executive function. The on-screen instructions read: “Is this a target card?”. A playing card is shown face up in the centre of the screen with the word “Number” or “Color” above it. If “Color” the participant is instructed to guess whether the target card is black or red. If the word is “Number” the participant must guess whether the current number displayed on the card is correct. At the start of the task, the participant guesses. Feedback on whether their answers are correct or not is provided. Subsequently, the participant may not proceed to the next trial until a correct response has been made. Part-way through the task the hidden rule changes. The participant is not told when these set-shifts occur, and they must learn the new target rule to proceed through the test. The participant is encouraged to work as quickly and accurately as possible.

### Technology use questionnaire

**S1 Fig.**
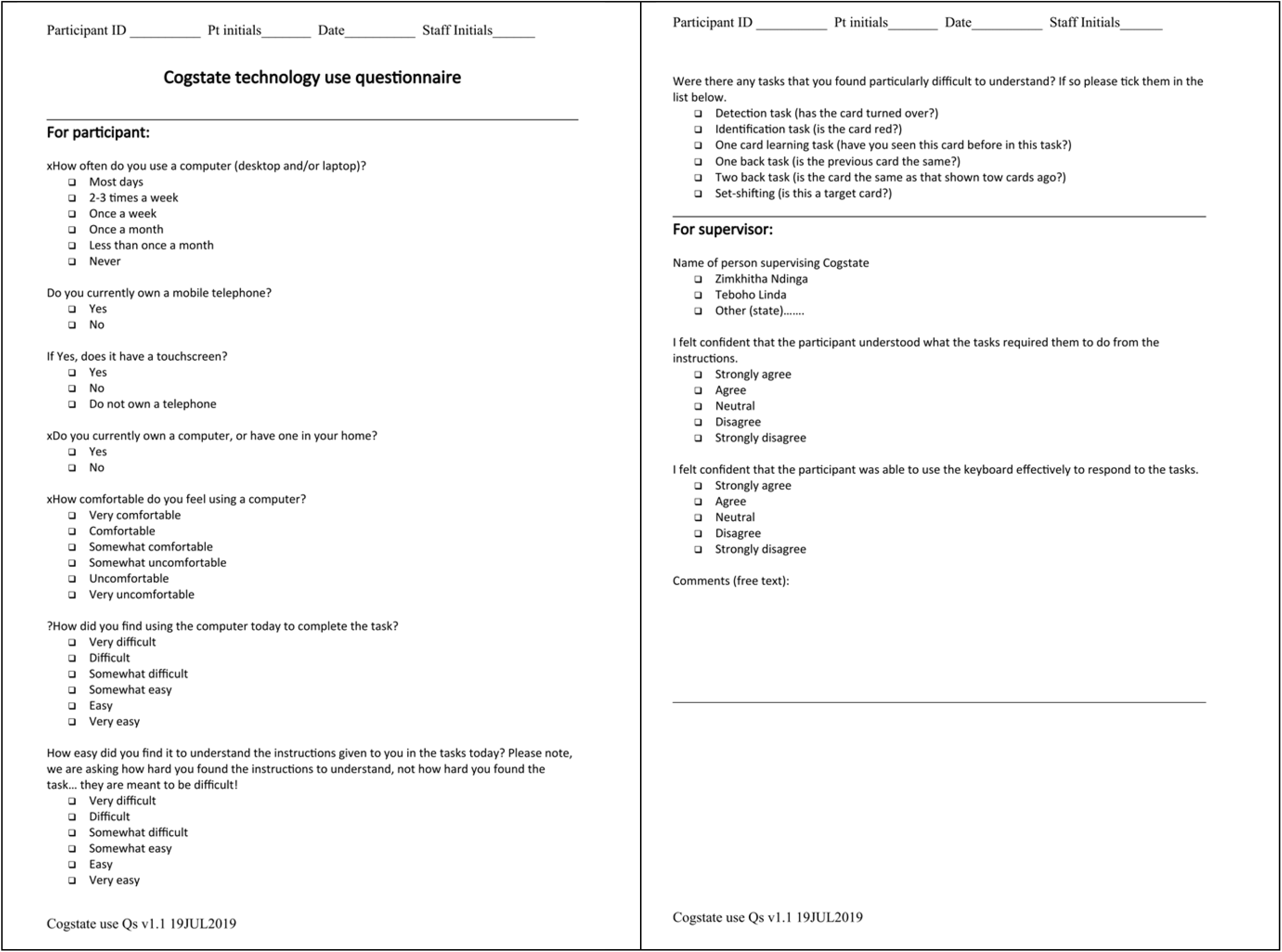
Technology use questionnaire used in CONNECT study.

## Results

**S2 Fig.**
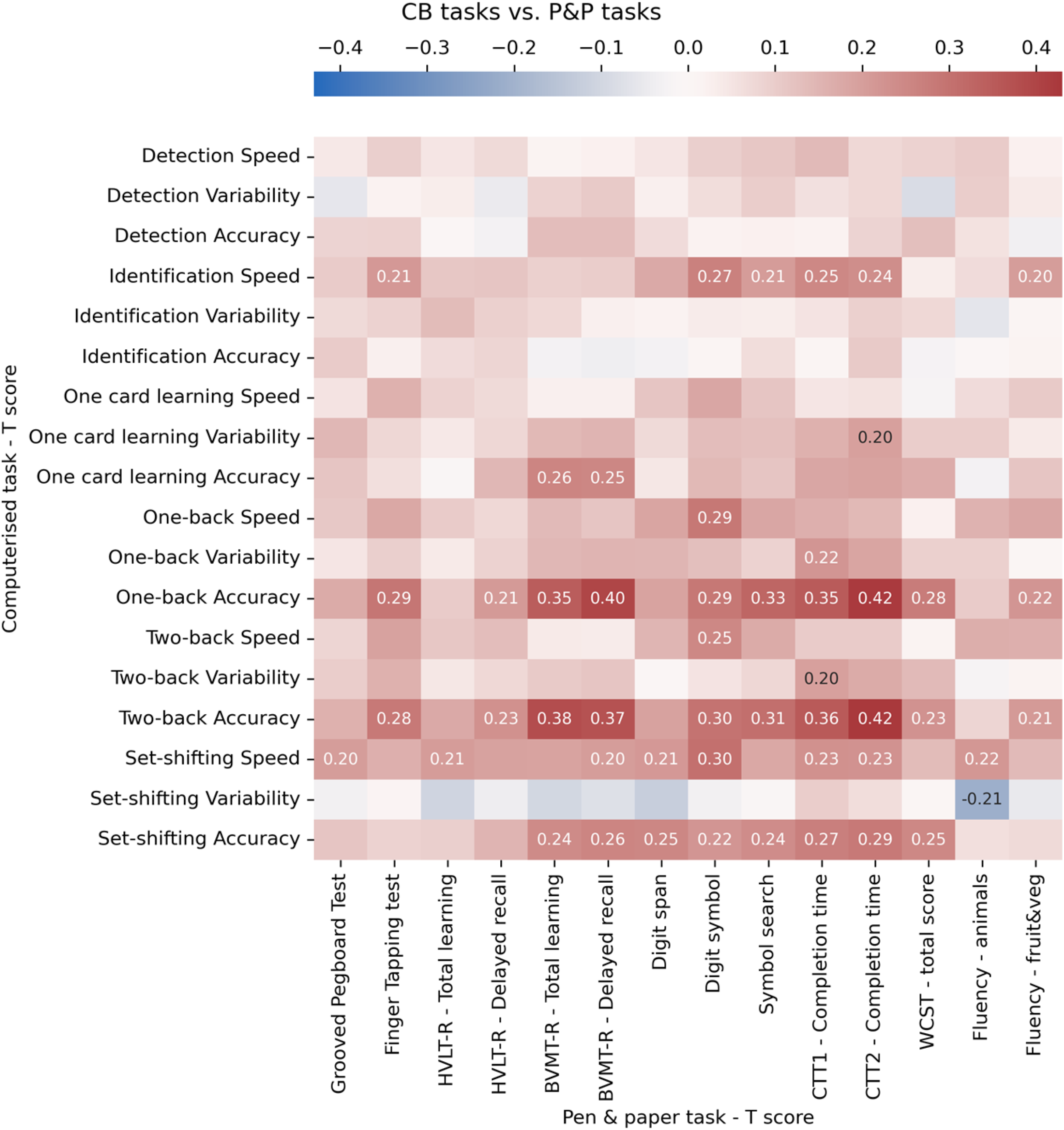
Heatmap of Pearson correlation coefficients between CB tasks and P&P tasks.

**S3 Fig.**
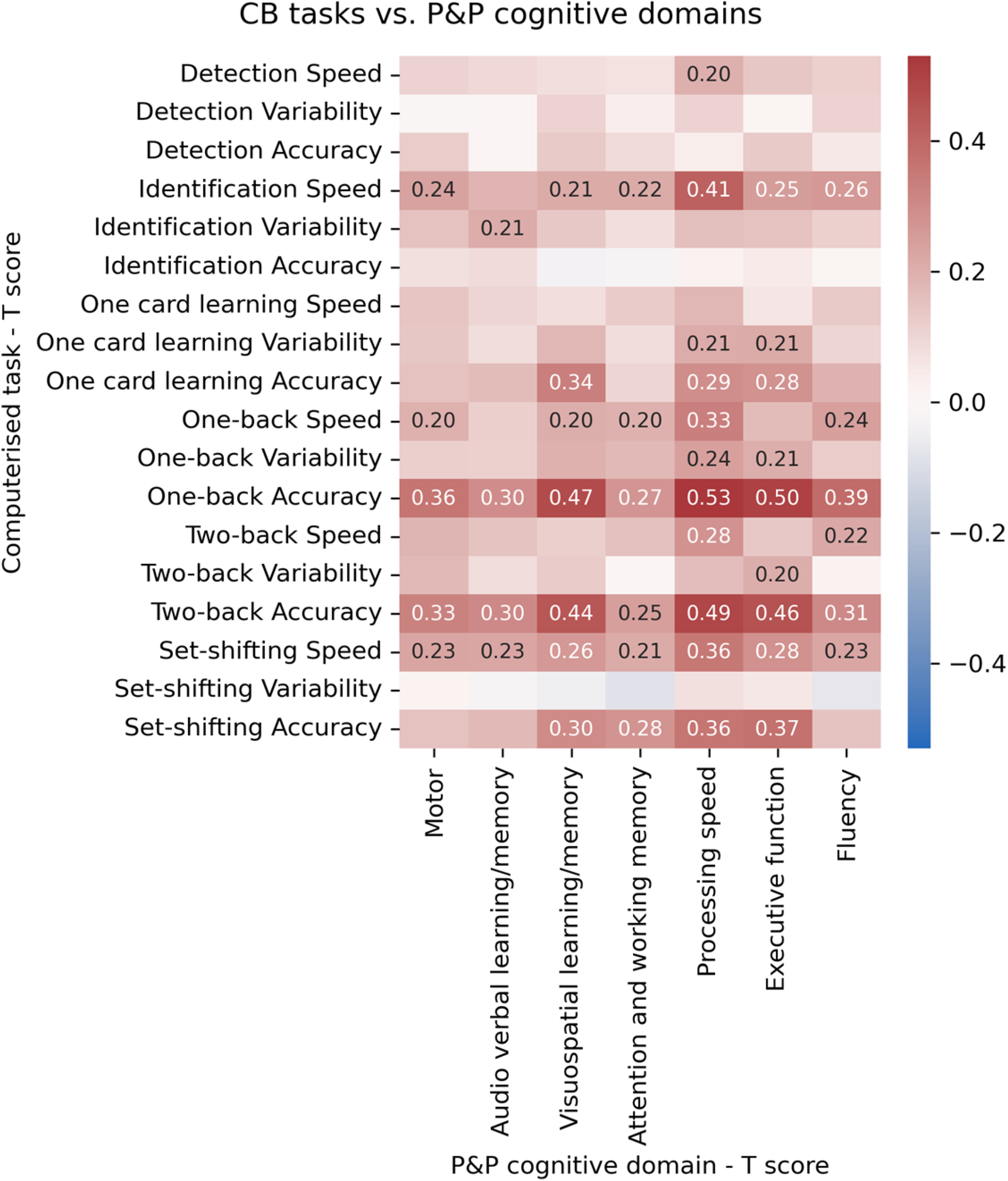
Heatmap of Pearson correlation coefficients between CB outcome measures and P&P domains.

